# FACTORS ASSOCIATED WITH MALARIA CASES AMOUNG ADULT PATIENTS AT CHONGWE DISTRICT HOSPITAL, ZAMBIA

**DOI:** 10.64898/2026.01.05.26343422

**Authors:** Mukuka Nkole Mulenga, Mukumbuta Nawa, Estella Sinkala, Priscilla Funduluka

## Abstract

**Introduction:** Malaria research in Zambia has largely focused on pregnant women and children under five, yet adults also play a critical role in sustaining transmission. This study assessed the prevalence and factors associated with malaria among adults aged ≥18 years attending Chongwe District Hospital.

**Methodology:** The study used an analytical cross-sectional study design. Data was collected using a structured questionnaire capturing socio-demographic information, including age, sex, education level, occupation, and socio-economic status. Environmental and behavioural factors, such as water source and use of insecticide-treated nets (ITNs), were also assessed. Malaria infection was tested using rapid diagnostic tests (RDTs). Data was analysed using SPSS version 26, it was summarised using frequencies and percentages. Further, hypothesis tests such as Chi-square and Fisher’s exact test were done on categorical variables and multivariable logistic regression was used to test for associations.

**Results:** A total of 134 participants were enrolled in the study, and consisted of 73 females (54.5%) and 61 males (45.4%). The study revealed an adult malaria prevalence of 14.2% among those who visited the health facility. Those with low education were associated with increased odds (aOR 5.55, P-value 0.048) of malaria compared to those with secondary education. Other variables such as gender, age, occupation, use of ITNs and water source were not statistically significant.

**Conclusion:** This study found that the prevalence of malaria among adults attending Chongwe district hospital for various ailments was higher than the expected prevalence of malaria in Lusaka district. The study further found that malaria was higher among those with lower education compared to those with higher education.

## Introduction

Malaria research tends to be targeted at pregnant women and under-five children who experience the severe forms of the disease and deaths, however, in its transmission cycle, adults including males also play a significant role (Bansal and Kumar, 2018; Bogale et al., 2025; Okova et al., 2024). Adult males and adult non-pregnant females living in endemic areas tend to develop partial immunity to the disease and tend to fend off the establishment of symptomatic disease or experience mild forms of the disease, thus do not seek treatment and remain chronic carriers of parasites and transmit them to vector mosquitoes when bitten (Faraglia et al., 2025; Lata et al., 2024). Studies on chronic carriers of malaria are few but an emerging area of interest in the transmission dynamics (Li et al., 2024). In one cohort study in Senegal, older individuals aged 15 – 49 were found to have a higher burden of malaria infections which was atypical or asymptomatic during the wet season compared to children below the age of 10 years (Legendre et al., 2025). One study in Zambia in two rural districts which implemented both household screening and routine passive health facility detection found that the majority of the malaria infections (56.6%) detected over the study period were coming from household screening visits implying that most malaria chronic carriers do not seek treatment at health facilities (Hamainza et al., 2014). The above evidence therefore suggests that malaria in adults is an important component in the malaria transmission, however, due to reduced morbidity and mortality in adults, the area is not prioritised.

Studies such as the Malaria Indicator Surveys in Zambia and elsewhere do not test for malaria prevalence among adult population making the prevalence in the adult population not readily available. One study in Ethiopia which compared malaria prevalence among asymptomatic people in different age-groups found that malaria prevalence was 7.5% among under five children, 9.5% among those aged five to fourteen years and 4.0% among those aged above fifteen years (Dabaro et al., 2023). Whilst the prevalence in this study showed that malaria was higher in those aged 5 – 14 years, the prevalence was not statistically significant in all the age groups (P-value = 0.782) underscoring the fact that malaria prevalence in older children or adults may not be significantly different from under-five children even though it is not regularly measured (Dabaro et al., 2023). This study therefore assessed the prevalence and factors associated with malaria among adults aged 18 years and above accessing health services at Chongwe district hospital. The findings of the study will add to the body of knowledge on the prevalence of malaria in the adult population which is not often measured in national surveys. Further, national authorities can develop further studies on malaria among the adult populations in Zambia and similar settings where malaria is endemic.

## Materials and Methods

### Study design

This was an analytical cross-sectional study which was conducted over a period of three (3) months i.e., from February to April 2024 during the peak period of malaria transmission.

### Study setting

The study was conducted at Chongwe District Hospital. The hospital is located in Chongwe District, in Lusaka Province, Zambia. The hospital provides medical care services to a population of 313,389 people as of the 2022 Zambian Census. The community’s primary occupations include farming and trade which align with malaria risk factors.

### Study Population

The study population included all patients aged 18 years and above who sought medical services at Chongwe District Hospital during the study period, regardless of malaria symptoms or not at the time of the study.

### Inclusion Criteria

- The study included all local residents who were aged 18 years or older who presented at Chongwe district hospital during the study period irrespective of malaria symptoms.
- People who have resided in Chongwe district for at least three months.

### Exclusion Criteria

- Patients in critical health conditions,
- individuals with mental health disorders.

#### 5.1.1 Sample size and Power

The sample size of the study was determined using the population proportion formula at 95% and alpha at 5%.

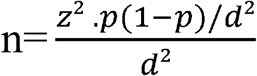

where n is the sample size, z is the z statistic for a level of confidence (z = 1.96 at 95% CI), d is the precision (if 5%, d = 0.05), and p is the proportion of malaria prevalence (p = 9.7%) from a malaria infection prevalence in a study by (Eisele et al., 2011) in Luangwa District.

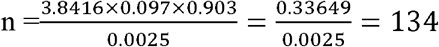

### Data collection

A standard structured questionnaire was used to collect data on sociodemographic characteristics and malaria infection was tested in the laboratory for each recruited participant using Rapid Diagnostic Tests (RDTs).

### 5.1 Data management and statistical analysis

#### Statistical Analysis Plan

Data were analysed using SPSS version 26. Descriptive statistics (frequencies and percentages) summarized participant characteristics. Associations between malaria infection and independent variables were assessed using Binary logistic regression. Variables with p < 0.2 in bivariate analysis were entered into multivariable logistic regression. Adjusted odds ratios (aORs) with 95% confidence intervals were reported, and p < 0.05 was considered statistically significant. Model fit was assessed using Akaike Information Criterion (AIC) and Bayesian Information Criterion (BIC).

### 5.2 Ethical Considerations

Written Informed consent was obtained from participants. Ethical approval was obtained from the ERES Converge Ethics Committee (Ref. No: [2024-Apr-006]) and the Chongwe District Health Office. Written informed consent was obtained from all participants prior to enrolment. Privacy of each study participant’s information was observed in that all the information recorded was kept under lock and key only accessible to the principal investigator. All clinical results were only made available to the clinicians for further treatment. Anonymity and confidentiality were observed at all times using coded identity numbers.

## Results

The study enrolled 134 participants, all of whom completed the questionnaire in line with the inclusion and exclusion criteria. They consisted of 73 females (54.5%) and 61 males (45.4%), with an equal distribution of marital status—50% married and 50% single. Participants with a secondary level of education formed the largest group, comprising 66 individuals (49.3%). Farmers represented the highest occupational group at 39 participants (29.1%), while security personnel were the least represented with just 1 participant (0.7%). Table 2 summarises the basic characteristics of the participants.

**Table 1:** Basic Characteristics of Participants.

| *Variable* |  | *N* | *Percentage* |
| --- | --- | --- | --- |
| **Gender** | *Male* | 61 | 45.5% |
|  | *Female* | 73 | 54.5% |
| **Marital status** | *Married* | 67 | 50.0% |
|  | *Single* | 67 | 50.0% |
| **Level of Education** | *Secondary* | 66 | 49.3% |
|  | *Tertiary* | 51 | 38.1% |
|  | *Primary* | 17 | 12.7% |
| **Occupation** | *Farmer* | 39 | 29.1% |
|  | *Teacher* | 12 | 9.0% |
|  | *Engineer* | 2 | 1.5% |
|  | *Self employed* | 33 | 24.7% |
|  | *House wife* | 6 | 4.5% |
|  | *Student* | 13 | 9.7% |
|  | *Medical* | 22 | 16.4% |
|  | *Unemployed* | 6 | 4.5% |
|  | *Security* | 1 | 0.7% |
| **Age** | *31-45yrs* | 44 | 32.8% |
|  | *18-30yrs* | 70 | 52.2% |
|  | *Above 45yrs* | 20 | 14.9% |
| **Use of ITNs** | *No* | 84 | 62.7% |
|  | *Yes* | 50 | 37.3% |
| **Are ITNs readily accessible** | *No* | 63 | 47.0% |
|  | *Yes* | 71 | 53.0% |
| **Water source** | *Bore-holed water* | 86 | 64.2% |
|  | *Tap water* | 48 | 35.8% |
| **Malaria infection season** | *Rainy season* | 78 | 58.2% |
|  | *Dry season* | 48 | 35.8% |
|  | *Cold season* | 8 | 6.0% |

**Table 2:**
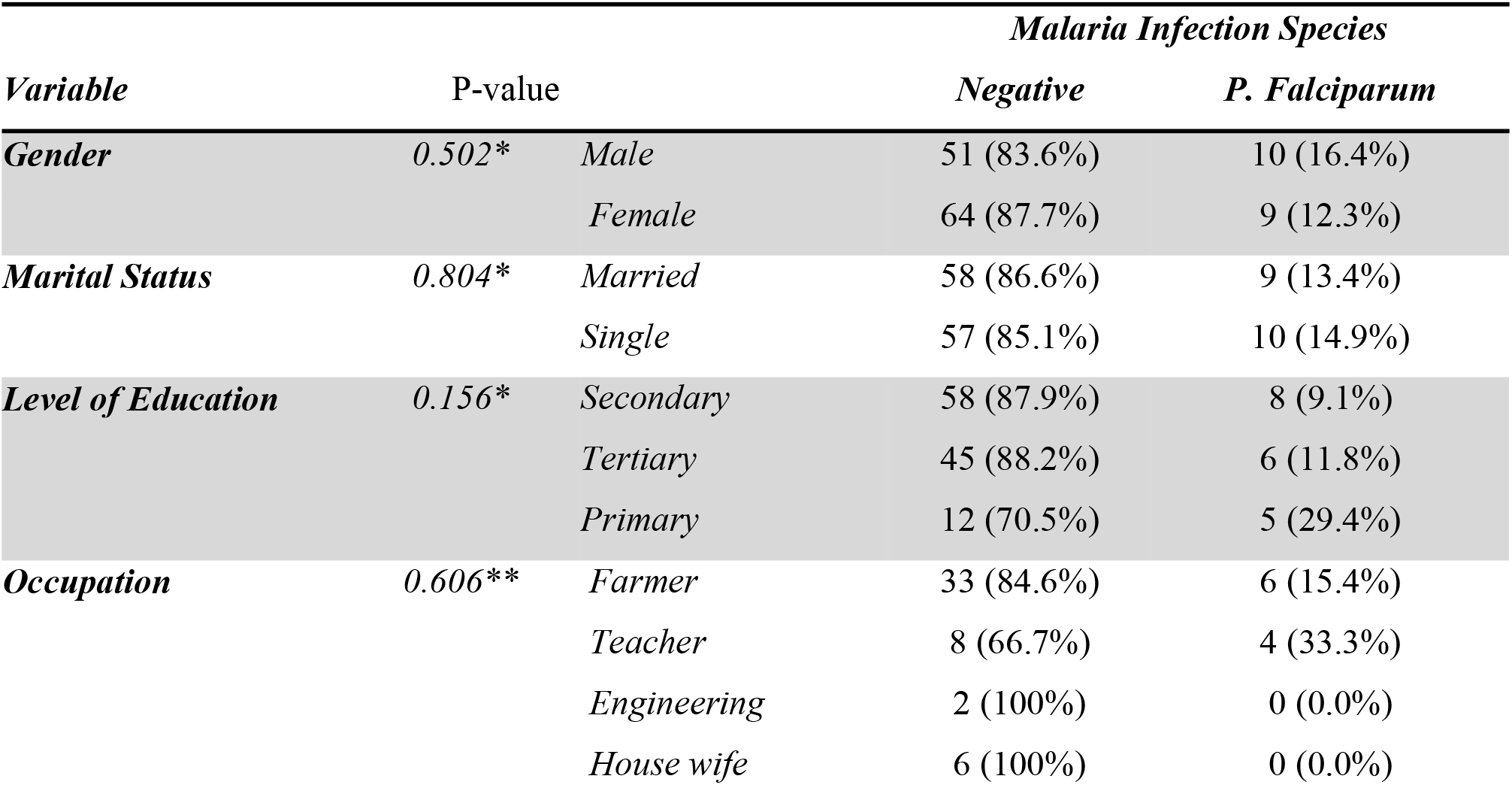
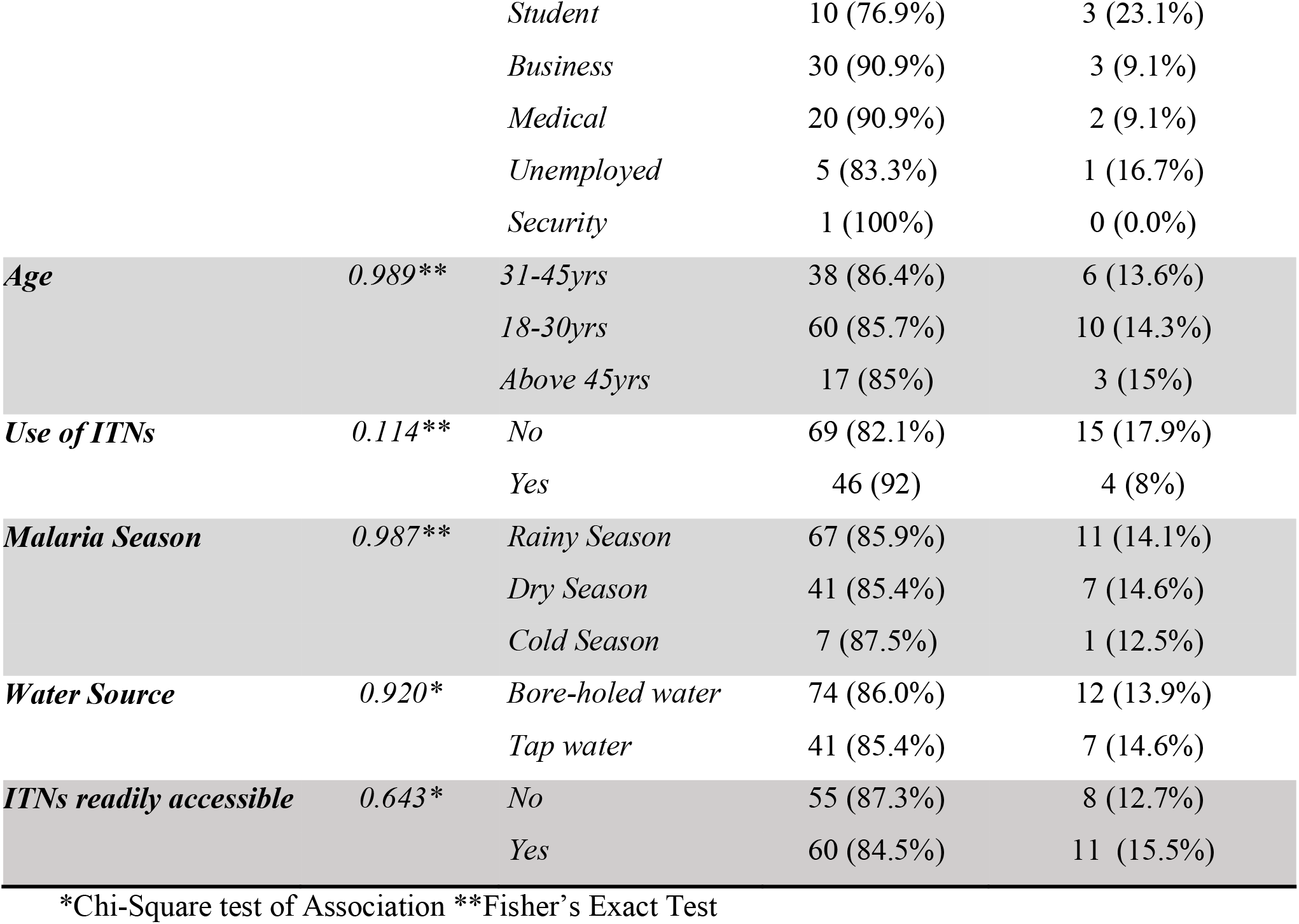
Distribution and Association of Demographic factors with Malaria Infection among demographic Factors.

| *Variable* | *P-value* |  | *Malaria Infection Species* |  |
| --- | --- | --- | --- | --- |
|  |  |  | *Negative* | *P. Falciparum* |
| **Gender** | 0.502* | *Male* | 51 (83.6%) | 10 (16.4%) |
|  |  | *Female* | 64 (87.7%) | 9 (12.3%) |
| **Marital Status** | 0.804* | *Married* | 58 (86.6%) | 9 (13.4%) |
|  |  | *Single* | 57 (85.1%) | 10 (14.9%) |
| **Level of Education** | 0.156* | *Secondary* | 58 (87.9%) | 8 (9.1%) |
|  |  | *Tertiary* | 45 (88.2%) | 6 (11.8%) |
|  |  | *Primary* | 12 (70.5%) | 5 (29.4%) |
| **Occupation** | 0.606** | *Farmer* | 33 (84.6%) | 6 (15.4%) |
|  |  | *Teacher* | 8 (66.7%) | 4 (33.3%) |
|  |  | *Engineering* | 2 (100%) | 0 (0.0%) |
|  |  | *House wife* | 6 (100%) | 0 (0.0%) |
|  |  | *Student* | 10 (76.9%) | 3 (23.1%) |
|  |  | *Business* | 30 (90.9%) | 3 (9.1%) |
|  |  | *Medical* | 20 (90.9%) | 2 (9.1%) |
|  |  | *Unemployed* | 5 (83.3%) | 1 (16.7%) |
|  |  | *Security* | 1 (100%) | 0 (0.0%) |
| **Age** | 0.989** | *31-45yrs* | 38 (86.4%) | 6 (13.6%) |
|  |  | *18-30yrs* | 60 (85.7%) | 10 (14.3%) |
|  |  | *Above 45yrs* | 17 (85%) | 3 (15%) |
| **Use of ITNs** | 0.114** | *No* | 69 (82.1%) | 15 (17.9%) |
|  |  | *Yes* | 46 (92) | 4 (8%) |
| **Malaria Season** | 0.987** | *Rainy Season* | 67 (85.9%) | 11 (14.1%) |
|  |  | *Dry Season* | 41 (85.4%) | 7 (14.6%) |
|  |  | *Cold Season* | 7 (87.5%) | 1 (12.5%) |
| **Water Source** | 0.920* | *Bore-holed water* | 74 (86.0%) | 12 (13.9%) |
|  |  | *Tap water* | 41 (85.4%) | 7 (14.6%) |
| **ITNs readily accessible** | 0.643* | *No* | 55 (87.3%) | 8 (12.7%) |
|  |  | *Yes* | 60 (84.5%) | 11 (15.5%) |

\*Chi-Square test of Association \*\*Fisher's Exact Test

**Table 3:**
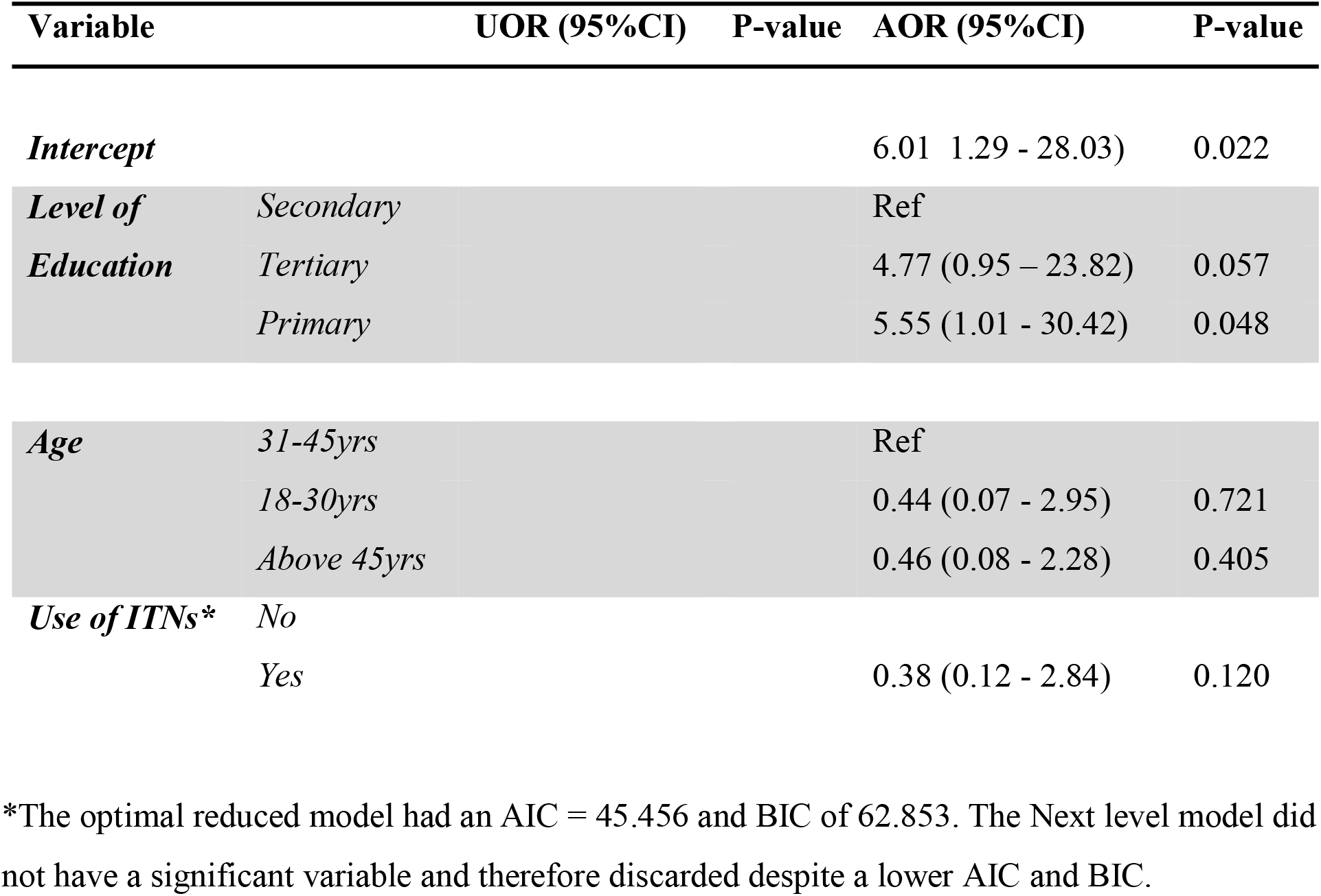
Reduced Model of Factors Associated with Malaria Infection.

### 6.2. The Distribution of Malaria Infection and Association of Demographic factors

The study found an overall prevalence of malaria of 14.2% (19/134) out of which ten (16.4%) were males and nine (12.3%) were females, however, the difference in the male/female prevalence was not statistically significant (P-value = 0.502). Similarly, there were no statistically significant differences (P value = 0.802) between among those infected with malaria between those who were married (13.4%) and those who were single (14.9%).

#### Multivariable Logistic Regression and Development of the Reduced Model

In the initial full multivariable logistic regression model, several demographic and behavioural variables were entered based on their bivariate associations. However, most predictors, including gender, marital status, occupation, age, malaria season, and water source, did not reach statistical significance and contributed to poor model fit, with wide confidence intervals and unstable estimates Akaike and Bayesian Information (AIC = 124.606, BIC = 127.546). Primary education emerged as a strong predictor of malaria infection, with markedly increased odds (OR = 27.21, 95% CI: 1.81–409.94, p = 0.017), while tertiary education suggested elevated risk but was not statistically significant (AOR = 5.35, p = 0.065). Use of insecticide-treated nets (ITNs) demonstrated a protective trend (AOR = 0.27, p = 0.066), though not significant in the adjusted model. To refine the analysis and improve stability, a stepwise backward reduction was applied.

The reduced model retained education level, age group, and ITN use, resulting in improved fit statistics (AIC = 45.456, BIC = 62.853).

In this optimized model, primary education remained significantly associated with malaria infection (AOR = 5.55, 95% CI: 1.01–30.42, p = 0.048), while tertiary education (AOR = 4.77, p = 0.057) and ITN use (AOR = 0.38, p = 0.120) showed important but non-significant trends.

## Discussion

In the present study, a malaria prevalence of 14.2% in adults was observed, which shows a higher than expected prevalence of malaria in Chongwe district of Lusaka province which has historically has been considered as having low to very low malaria prevalence rates of below 5% (Lowa et al., 2018; Nawa et al., 2024). In this study, malaria prevalence may have been higher than expected because the study was a facility based study which only included people who sought health services, however, the people had different ailments therefore not everyone was symptomatic for malaria (Asmelash et al., 2025; Mesele et al., 2025; Yutura et al., 2024). The authors did not find any recent studies on malaria prevalence in Chongwe district, however, one old study had found a prevalence of 45% among household contacts of passively detected cases at a local health facility in Chongwe (Pinchoff et al., 2015). The referenced study found a higher prevalence because it only enrolled people from households where a malaria case already existed and therefore did not necessarily measure population level malaria prevalence (Pinchoff et al., 2015). This study therefore provides evidence of malaria prevalence among adults attending a health facility in Chongwe district who are not traditionally assessed for malaria during routine national surveys such as the Malaria Indicator Surveys (MIS). The authors therefore recommend future studies to be done using a random household survey among the adult population in Chongwe and other low transmission settings to further assess the malaria prevalence in the adult population as nationwide malaria classification by endemicity is based on measurements in under-five children (Nawa et al., 2024).

Furthermore, this study demonstrated that participants with primary education had significantly higher odds (aOR = 5.55), of malaria infection compared to those with secondary education reinforcing the association between lower education levels and vulnerability to malaria (Alao et al., 2025; Nzoputam et al., 2024). This finding aligns with a systematic review that included 75 studies from sub-Saharan Africa which found that those with higher education levels were less likely to have malaria compared to those with lower educational attainments (Degarege et al., 2019). This can be explained by the fact that those with lower educational attainments were less likely to use preventive measures such as insecticide treated nets (Clouston et al., 2015; Firdaus et al., 2025). One study in Zambia showed that people who have lower education were less likely to use malaria prevention interventions such as insecticide treated nets (ITNs) (Jumbam et al., 2020). This was also supported by other studies elsewhere that people with low educational attainments were less likely to use preventive measures against malaria compared to those with higher education (Kouamé et al., 2022; Mbishi et al., 2024; Nzoputam et al., 2024). This study therefore underscores the need for health education in Chongwe district especially targeting people with low education in the communities (Onyinyechi et al., 2024).

The main limitation of this study was that it was a health facility-based study, as such the prevalence that was found did not represent the general population but only adults that seek health services from the district hospital in Chongwe.

## Conclusion

This study found that the prevalence of malaria among adults attending Chongwe District Hospital for various ailments was higher than the expected prevalence of malaria in Lusaka province. The study further found that malaria was higher among those with lower education compared to those with higher education. The study therefore recommends addressing malaria among adults in malaria hotspot areas such as Chongwe district in Lusaka province which has a low malaria prevalence. Further, it recommends targeted interventions for vulnerable populations such as those with low education.

## Data Availability

All data produced in the present study are available upon reasonable request to the authors

## REFERENCES

Alao, J.O., Olowoshile, O.P., James, T.A., Okezie, C.C., Adebayo, Z.P., Ogbonna, S.C., Oyelayo, E.A., 2025. Socioeconomic and educational influences on malaria prevention and treatment behaviours in rural Nigeria. BMC Public Health 25, 3079. 10.1186/s12889-025-24326-3

Asmelash, D., Agegnehu, W., Fenta, W., Asmelash, Y., Debebe, S., Asres, A., 2025. The Burden of Asymptomatic Malaria Infection in Children in Sub-Saharan Africa: A Systematic Review and Meta-Analysis Exploring Barriers to Elimination and Prevention. J. Epidemiol. Glob. Health 15, 17. 10.1007/s44197-025-00365-2

Bansal, G.P., Kumar, N., 2018. Immune Responses in Malaria Transmission. Curr. Clin. Microbiol. Rep. 5, 38–44. 10.1007/s40588-018-0078-x

Bogale, K.A., Yenesew, M.A., Alemu, K., Muchie, K.F., Asemahagn, M.A., Enbiale, W., 2025. Facilitators and barriers of malaria prevention and treatment services to pregnant women in Ethiopia: a multi-level health system analysis, 2025. Malar. J. 24, 337. 10.1186/s12936-025-05555-8

Clouston, S.A.P., Yukich, J., Anglewicz, P., 2015. Social inequalities in malaria knowledge, prevention and prevalence among children under 5 years old and women aged 15–49 in Madagascar. Malar. J. 14, 499. 10.1186/s12936-015-1010-y

Dabaro, D., Birhanu, Z., Adissu, W., Yilma, D., Yewhalaw, D., 2023. Prevalence and predictors of asymptomatic malaria infection in Boricha District, Sidama Region, Ethiopia: implications for elimination strategies. Malar. J. 22, 284. 10.1186/s12936-023-04722-z

Degarege, A., Fennie, K., Degarege, D., Chennupati, S., Madhivanan, P., 2019. Improving socioeconomic status may reduce the burden of malaria in sub Saharan Africa: A systematic review and meta-analysis. PLOS ONE 14, e0211205. 10.1371/journal.pone.0211205

Eisele, T.P., Miller, J.M., Moonga, H.B., Hamainza, B., Hutchinson, P., Keating, J., 2011. Malaria Infection and Anemia Prevalence in Zambia’s Luangwa DistrictLJ: An Area of Near-Universal Insecticide-Treated Mosquito Net Coverage 84, 152–157. 10.4269/ajtmh.2011.10-0287

Faraglia, F., Vita, S., Kontogiannis, D., Bartoli, T.A., Rosati, S., Bevilacqua, N., Maffongelli, G., Corpolongo, A., Bartolini, B., Vulcano, A., D’Abramo, A., Fontana, C., Nicastri, E., Mobile-INMI group, 2025. Burden of asymptomatic malaria in adult sub-Saharan migrants attending an outpatient clinic in Rome from February 2024 to January 2025. Infect. Dis. Poverty 14, 107. 10.1186/s40249-025-01379-5

Firdaus, M.H., Wan Puteh, S.E., Sutan, R., Abdul Manaf, M.R., 2025. Effectiveness of family health education in malaria elimination programmes: a scoping review. Malar. J. 24, 144. 10.1186/s12936-025-05371-0

Hamainza, B., Moonga, H., Sikaala, C.H., Kamuliwo, M., Bennett, A., Eisele, T.P., Miller, J., Seyoum, A., Killeen, G.F., 2014. Monitoring, characterization and control of chronic, symptomatic malaria infections in rural Zambia through monthly household visits by paid community health workers. Malar. J. 13, 128. 10.1186/1475-2875-13-128

Jumbam, D.T., Stevenson, J.C., Matoba, J., Grieco, J.P., Ahern, L.N., Hamainza, B., Sikaala, C.H., Chanda-Kapata, P., Cardol, E.I., Munachoonga, P., Achee, N.L., 2020. Knowledge, attitudes and practices assessment of malaria interventions in rural Zambia. BMC Public Health 20, 216. 10.1186/s12889-020-8235-6

Kouamé, R.M.A., Guglielmo, F., Abo, K., Ouattara, A.F., Chabi, J., Sedda, L., Donnelly, M.J., Edi, C., 2022. Education and Socio-economic status are key factors influencing use of insecticides and malaria knowledge in rural farmers in Southern Côte d’Ivoire. BMC Public Health 22, 2443. 10.1186/s12889-022-14446-5

Lata, S., Gupta, S.K., Kumar, G., Yadav, S., Mohanty, S.S., Prasad, P., Singh, B., Singh, S., Saroha, P., Kumar, D., Singh, P., Vikram, K., Savargaonkar, D., Singh, H., 2024. Moving population is a challenge for malaria elimination in India: A cross-sectional study to assess malaria parasite infections in walking pilgrims in western Rajasthan, India. IJID Reg. 12, 100418. 10.1016/j.ijregi.2024.100418

Legendre, E., Ciré Ba, E.-H.K., L’Ollivier, C., Cissoko, M., Katile, A., Mehadji, M., Serre, P., Sokhna, C., Ranque, S., Danfakha, F., Bendiane, M.-K., Sagara, I., Gaudart, J., Landier, J., 2025. Plasmodium falciparum carriage in a population under long-term, intensive malaria control in Kedougou region, Senegal: a 1-year cohort study. Lancet Glob. Health 13, e1935–e1945. 10.1016/S2214-109X(25)00271-2

Li, J., Docile, H.J., Fisher, D., Pronyuk, K., Zhao, L., 2024. Current Status of Malaria Control and Elimination in Africa: Epidemiology, Diagnosis, Treatment, Progress and Challenges. J. Epidemiol. Glob. Health 14, 561–579. 10.1007/s44197-024-00228-2

Lowa, M., Sitali, L., Siame, M., Musonda, P., 2018. Human mobility and factors associated with malaria importation in Lusaka district, Zambia: a descriptive cross sectional study. Malar. J. 17, 404. 10.1186/s12936-018-2554-4

Mbishi, J.V., Chombo, S., Luoga, P., Omary, H.J., Paulo, H.A., Andrew, J., Addo, I.Y., 2024. Malaria in under-five children: prevalence and multi-factor analysis of high-risk African countries. BMC Public Health 24, 1687. 10.1186/s12889-024-19206-1

Mesele, F.B., Belay, D.S., Gebreslassie, K.B., Abrha, M.G., Tesfay, B.G., Gebreanenia, F.A., 2025. Prevalence of malaria and its associated factors among adults living in Samre Woreda, Tigray, Ethiopia, 2023/24: a community-based cross-sectional study. BMC Public Health 25, 2162. 10.1186/s12889-025-23423-7

Nawa, M., Mupeyo-Mudala, C., Banda-Tembo, S., Adetokunboh, O., 2024. The effects of modern housing on malaria transmission in different endemic zones: a systematic review and meta-analysis. Malar. J. 23, 235. 10.1186/s12936-024-05059-x

Nzoputam, C.I., Ogidan, O.C., Barrow, A., Ekholuenetale, M., 2024. What do women in the highest malaria burden country know about ways to prevent malaria? A multi-level analysis of the 2021 Nigeria Malaria Indicator Survey data. Malar. J. 23, 361. 10.1186/s12936-024-05195-4

Okova, D., Lukwa, A.T., Oyando, R., Bodzo, P., Chiwire, P., Alaba, O.A., 2024. Malaria Prevention for Pregnant Women and Under-Five Children in 10 Sub-Saharan Africa Countries: Socioeconomic and Temporal Inequality Analysis. Int. J. Environ. Res. Public. Health 21, 1656. 10.3390/ijerph21121656

Onyinyechi, O.M., Ismail, S., Nashriq Mohd Nazan, A.I., 2024. Prevention of malaria in pregnancy through health education intervention programs on insecticide-treated nets use: a systematic review. BMC Public Health 24, 755. 10.1186/s12889-024-17650-7

Pinchoff, J., Henostroza, G., Carter, B.S., Roberts, S.T., Hatwiinda, S., Hamainza, B., Hawela, M., Curriero, F.C., 2015. Spatial patterns of incident malaria cases and their household contacts in a single clinic catchment area of Chongwe District, Zambia. Malar. J. 14, 305. 10.1186/s12936-015-0793-1

Yutura, G., Massebo, F., Eligo, N., Kochora, A., Wegayehu, T., 2024. Prevalence of malaria and associated risk factors among household members in South Ethiopia: a multi-site cross-sectional study. Malar. J. 23, 143. 10.1186/s12936-024-04965-4

